# Quality Control Metrics for Whole Blood Transcriptome Analysis in the Parkinson’s Progression Markers Initiative (PPMI)

**DOI:** 10.1101/2021.01.05.21249278

**Authors:** Elizabeth Hutchins, David Craig, Ivo Violich, Eric Alsop, Bradford Casey, Samantha Hutten, Alyssa Reimer, Timothy G. Whitsett, Karen L. Crawford, Arthur W Toga, Shawn Levy, Madison Robinson, Nripesh Prasad, J. Raphael Gibbs, Mark R. Cookson, Kendall Van Keuren-Jensen, Parkinson Progression Marker Initiative

## Abstract

The Michael J. Fox Foundation’s Parkinson’s Progression Markers Initiative (PPMI) is an observational study to comprehensively evaluate Parkinson’s disease (PD) patients using imaging, biologic sampling, clinical and behavioural assessments to identify biomarkers of PD progression. As part of this study, we obtained 4,756 whole blood samples from 1,570 subjects at baseline, 0.5, 1, 2, and 3 years from enrollment in the study. We isolated RNA and performed whole transcriptome sequencing in this longitudinal cohort. Here, we describe and quantify technical variability associated with this dataset through the use of pooled reference samples, including plate distribution, RNA quality, and outliers. This large, uniformly processed dataset is available to researchers at https://www.ppmi-info.org.

## Background & Summary

Parkinson’s disease (PD) is a chronic, degenerative neurological disorder with increasing incidence with age. As the population ages, PD is expected to impact more than 1.2 million people in the United States by 2030^1^. To date there are no effective or reliable biomarkers for PD diagnosis or progression. To help advance biomarker discovery, longitudinal studies with large sample sizes would improve our understanding of the genesis and progression of this disease. The Parkinson’s Progression Markers Initiative (PPMI) is aimed at identifying and validating biomarkers of PD progression by enrolling large numbers of participants across multiple sites and following them over years with both sample collection and deep clinical phenotyping (ClinicalTrials.gov Identifier: NCT01141023)^2^. Here, we describe a dataset using RNA-sequencing (RNA-seq) in the PPMI cohort using whole blood samples as a readily accessible peripheral biofluid.

As RNA is becoming a powerful tool in the development of biomarkers for the diagnosis and progression of disease^3^, the PPMI RNA-Sequencing Project includes longitudinal, whole blood sequencing data from 1,570 subjects at baseline, followed by 0.5-, 1-, 2- and 3-year visits. As the transcriptome changes with age and health status, analyses can reveal gene expression changes and pathways disrupted by disease processes. Analyses of readily accessible biofluids, such as blood, coupled with longitudinal collection, provide the potential to uncover valuable transcriptomic biomarkers. Additionally, the patients in the PPMI study have extensive clinical and imaging data, that is collected longitudinally^4^.

Whole-blood PAXgene RNA samples were collected from PPMI participants (Figure 1A) at multiple time points, including: 1) de novo PD subjects with a diagnosis of PD for two years or less and who are not taking PD medications at the time of enrollment (baseline), 2) unaffected subjects who are 30 years or older and who do not have a first degree blood relative with PD (control), 3) Subjects without evidence of a dopaminergic deficit (SWEDD), who were initially ascertained as PD subjects, but have a normal DaTscan, 4) prodromal, defined as people without a PD diagnosis but who have early comorbid conditions including hyposmia or REM sleep behaviour disorder (RBD), 5) genetic cohort, which includes both manifesting and non-manifesting carriers of mutations in the genes for leucine rich repeat kinase 2 (LRRK2), glucocerebrosidase beta (GBA), or alpha synuclein (SNCA)^5^, and 6) genetic registry subjects who have a genetic mutation in LRRK2, GBA, or SNCA or a first-degree relative with one of the above mutations (Table 1; ClinicalTrials.gov Identifier: NCT01141023). Clinical data collected on the cohort includes imaging (DaTscan), extensive questionnaires regarding medical and personal histories, along with cognitive and physical assessments, and whole genome sequencing data, all available at https://ppmi-info.org.

**Table 1.**
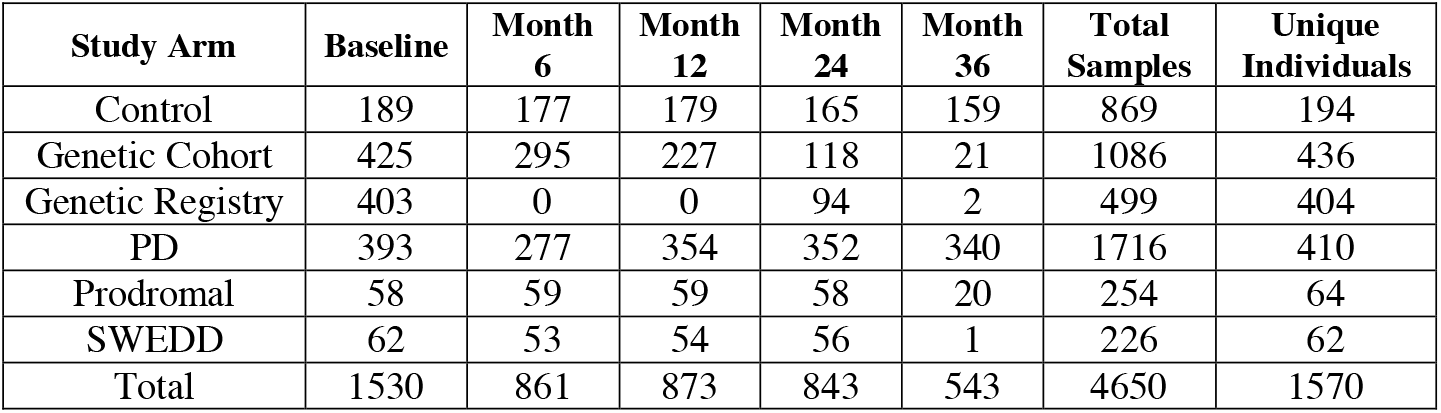
Sequenced samples by study arm and visit.

| Study Arm | Baseline | Month 6 | Month 12 | Month 24 | Month 36 | Total Samples | Unique Individuals |
| --- | --- | --- | --- | --- | --- | --- | --- |
| Control | 189 | 177 | 179 | 165 | 159 | 869 | 194 |
| Genetic Cohort | 425 | 295 | 227 | 118 | 21 | 1086 | 436 |
| Genetic Registry | 403 | 0 | 0 | 94 | 2 | 499 | 404 |
| PD | 393 | 277 | 354 | 352 | 340 | 1716 | 410 |
| Prodromal | 58 | 59 | 59 | 58 | 20 | 254 | 64 |
| SWEDD | 62 | 53 | 54 | 56 | 1 | 226 | 62 |
| Total | 1530 | 861 | 873 | 843 | 543 | 4650 | 1570 |

**Figure 1.**
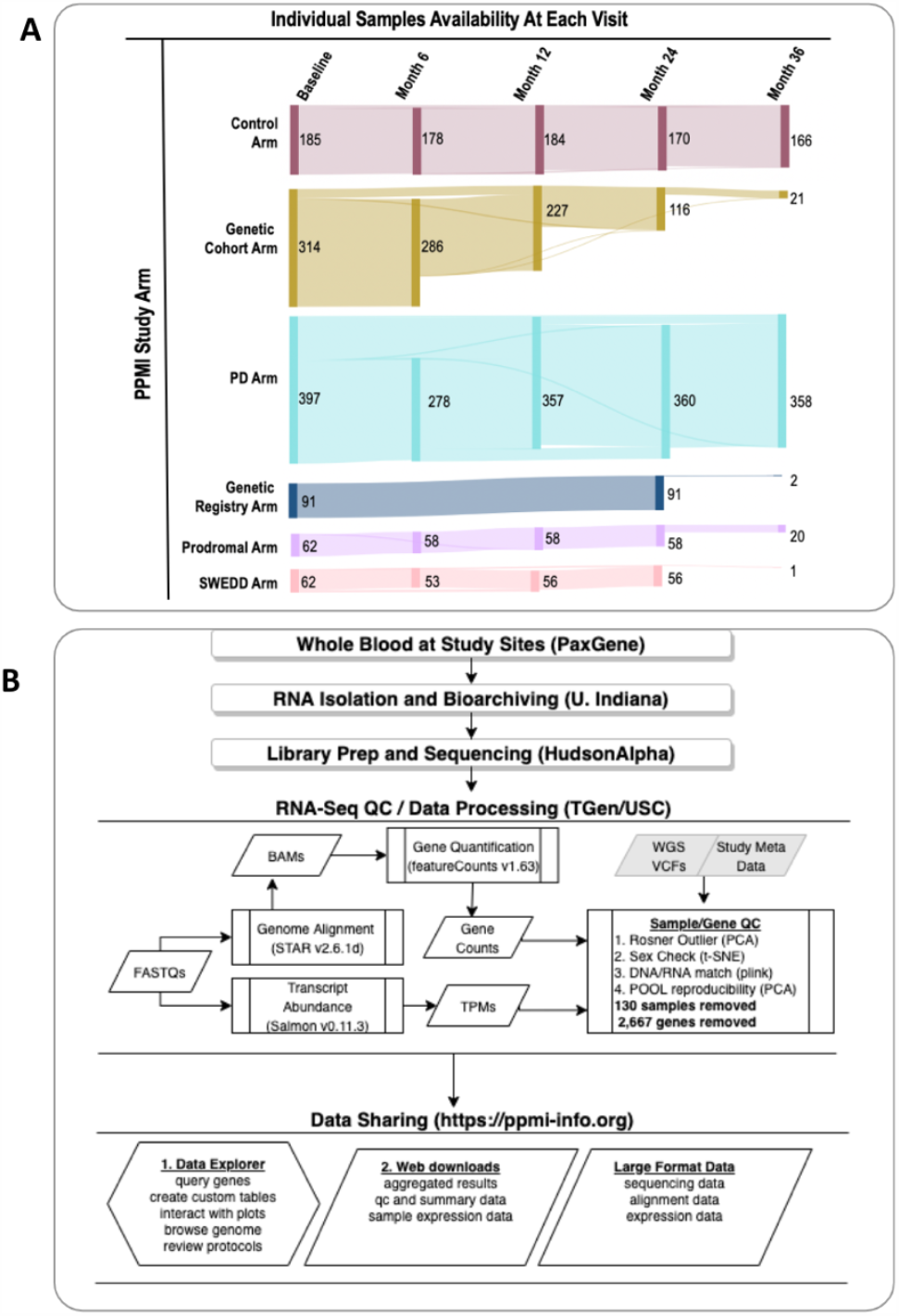
Sample and Data workflow. **(A)** Sankey diagram showing sample availability at each visit. The number of samples at each arm at baseline, month 6, month 12, month 24, and month 36. **(B)** Workflow of same sequencing, and initial data processing. Samples were sent to the biorepository at IU for RNA isolation, and then sent to HudsonAlpha Institute for Biotechnology for library preparation and sequencing. Genome alignment and gene and transcript quantification were performed (rectangles with end bars represent data analysis). Samples were assessed for outliers using a Rosner outlier test, sex incompatibility check, and a DNA/RNA incompatibility check. Pools were used to measure intra-plate variability of gene and transcript expression. FASTQs, BAMs, TPMs, and gene counts (rhomboids represent data type, while grey a background indicates data sourced from another portion of the PPMI study) are all available for download via the LONI IDA. A browser (a hexagon represents an interactive data explorer) at https://ppmi-info.org allows for querying of genes and exploration of the relationship to clinical parameters.

Here, we organized sequencing and data analysis pipelines in an effort to generate uniformly processed samples, from RNA isolation through gene and transcript detection (Figure 1B). Sequencing methods were specifically adopted to capture extensive transcriptomic information, and included steps for rRNA depletion, globin depletion and strandedness. The current study specifically describes sample quality metrics that were taken at multiple points throughout the process, that are reported here for researchers interested in using this cohort in additional analyses. We provide a table of the quality control metrics that were examined in this data, such as RNA Integrity Number (RIN), % useable bases, % rRNA, number of reads, insert size, number of genes detected, and other parameters (the metadata file is not included as it contains patient information. These data can be obtained through an application at: (https://amp-pd.org/). In addition, we described the use of technical replicates: aliquots from two, large uniform RNA pools that were placed on every plate in the study. We identified a series of inconsistent genes, due to technical variability across replicates, that are listed in Supplementary Table 1 for researchers to consider when analysing the data and interpreting results. These quality metrics, combined with additional information about the distribution of subject groups across plates will provide guidance to researchers in the effective design of projects employing this dataset.

## Methods

### Study Participants

Participants were enrolled at 33 clinical sites across the US (21), Europe (10), Israel (1), and Australia (1). The study was approved by the Research Subjects Review Board at the University of Rochester, and by the institutional review board at each site. Patients were divided into one of five study arms using information obtained upon enrollment (HC, PD, Genetic Registry, Genetic Cohort, and Prodromal) as described elsewhere^2^. Full inclusion and exclusion criteria are listed at clinicaltrials.gov (Identifier: <u>NCT01141023</u>) and at https://ppmi-info.org. The contribution of subjects from each site is shown in Figure 2A and their distribution across plates for sequencing can be visualized in Figure 2C. Study participants were added at different rates, and in Figure 2C, the irregular distribution of some cohorts and time points can be observed. For example, the 6-month samples are on the second half of the plates and the genetic registry and genetic cohort subjects are also predominantly in the second half of the plates.

**Figure 2.**
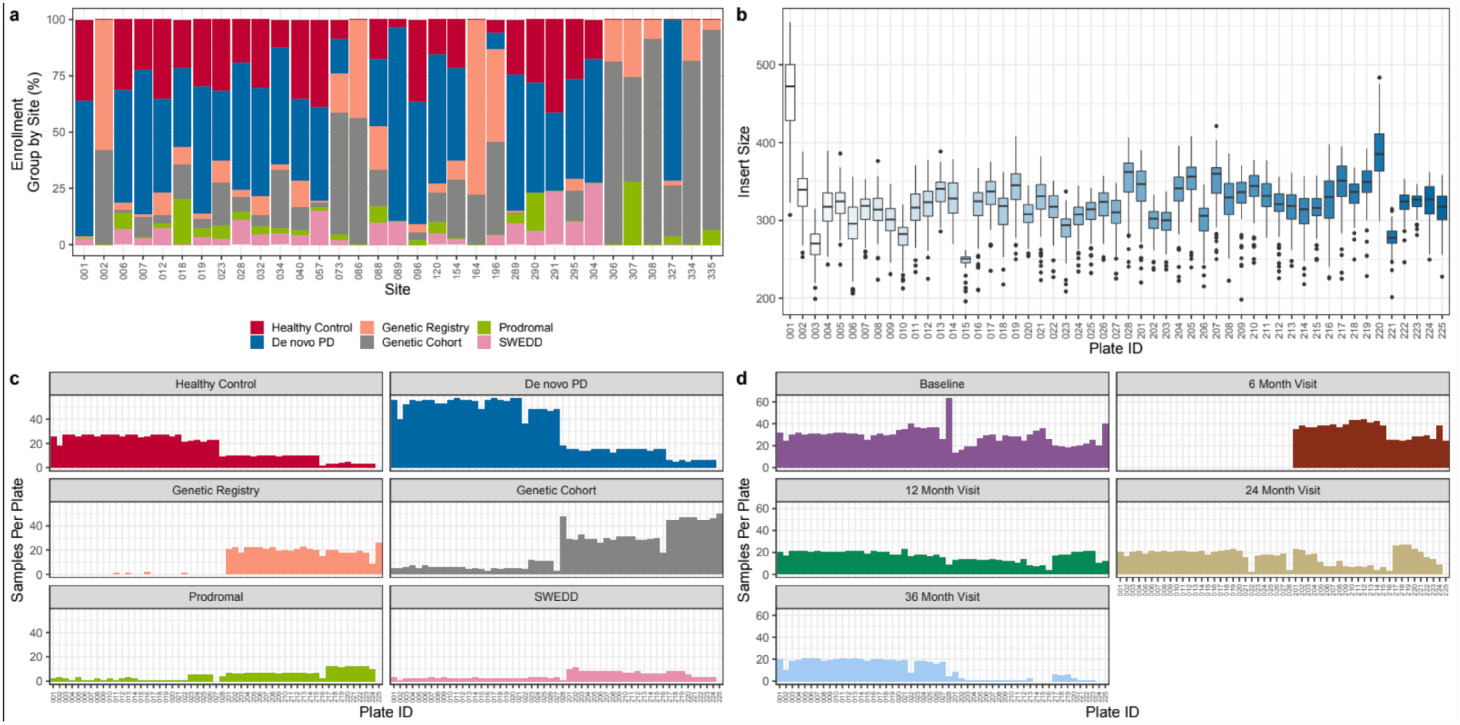
Sources of sample variability. **(a)** Different sites had varying proportions of study enrollment groups; sites 2, 73, 86, 164, 196, 306, 307, 308, 334, and 335 were mostly comprised of subjects in the Genetic Registry and Genetic Cohort. **(b)** Boxplot of the sample insert size by plate. Plate 1 is the clearest outlier, with a larger insert size. Plates 3 and plate 15 have smaller inserts. **(c, d)** Frequency plot showing the number of samples sequenced on each plate by **(c)** enrollment group and **(d)** visit.

### Sample Collection and RNA Isolation

The PPMI Biologics Manual, available at www.ppmi-info.org, gives a detailed description of the blood collection procedure. Between 8am and 10am, whole blood samples were collected from fasting individuals in PAXgene tubes (Qiagen). The subject followed a low-fat diet if fasting was not possible (information can be found at https://www.ppmi-info.org). PAXgene tubes were kept at room temperature for 24 hours before being frozen and shipped to the PPMI Biorepository Core at Indiana University (IU) within two weeks of collection. The PAXgene blood miRNA kit (Qiagen) was used to isolate total RNA, followed by DNase treatment. DNase-treated RNA samples were sent to the HudsonAlpha Institute of Biotechnology (HAIB) for library preparation and sequencing.

### Reference Pool Generation

In addition to subject samples, the PPMI Biorepository Core at IU created two RNA reference pools, one generated from RNA samples from PD subjects and the other from RNA samples from HC subjects. A total of 619 RNA aliquots from individuals with PD and 427 aliquots of whole blood RNA from HCs were thawed and combined to create two pools (PDPOOL and HCPOOL). Once pooled, 1 μg aliquots of the pool were distributed throughout the experiment so that each pool was run one time on each plate during sequencing, technical replicates. Sequencing performance of the pools are shown in the PCAs generated in Figure 3B and C.

**Figure 3.**
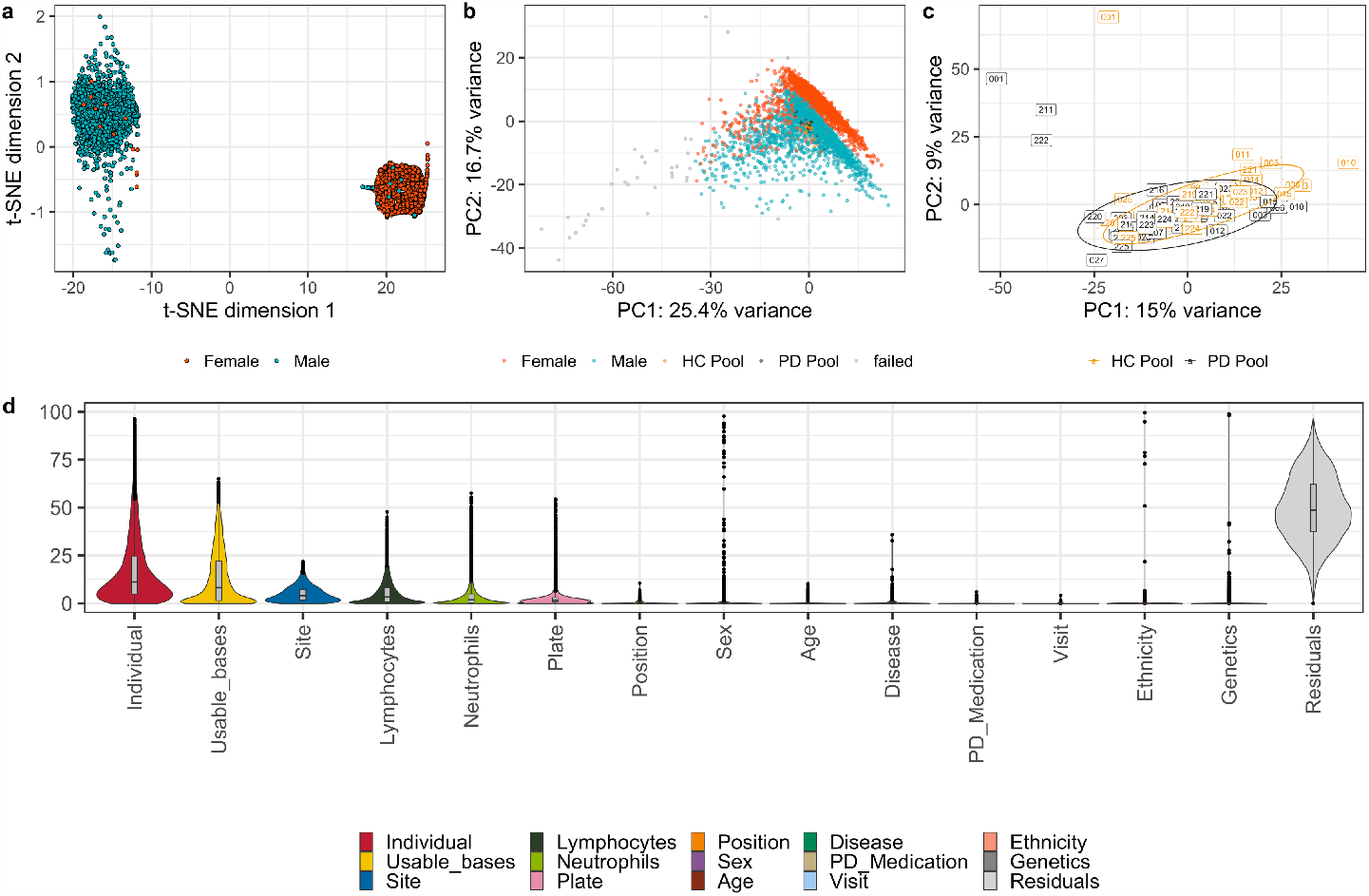
Intra-plate quality control. **(a)** t-NSE plot demonstrating the sex incompatibility check between clinically reported sex and genetically determined sex. **(b)** Principal component analysis (PCA) using the top 500 expressed genes labelled by clinically reported sex, HC and PD pool technical replicates, and samples that failed QC. **(c)** Principal component analysis (PCA) of the two pooled technical replicates sequenced on each plate (HC Pool and PD Pool). **(d)** Variance partition analysis at the gene-level after filtering out the most variable genes and transcripts. Primary sources of gene variation include individual subjects, usable bases, site, lymphocytes, and neutrophils. Primary sources of transcript variation include individual, usable bases, site, and blood chemistry.

### Library Preparation and Sequencing

Samples were processed in a 96-well format with a HC Pool and PD Pool sample placed on each plate. RNA was diluted to 30 ng/μl and 684-752 ng of RNA were used for Input for the rRNA and globin reduction steps. The remaining RNA was used in a separate study to assess the small RNA contents in the whole blood samples. The Illumina Globin-Zero Gold kit (catalog number GZG1224) was used to remove rRNA and globin. Stranded libraries were prepared by first-strand synthesis and second strand synthesis using the New England Biolabs (NEB) Ultra II First Strand Module (catalog number E7771L) followed by the NEB Ultra II Directional Second Strand Module (catalog number E7550L). cDNA was converted by standard, ligation-based library preparationr: 1) NEB End Repair Module (catalog number E6050L), 2) NEB A-tailing Module (catalog number E6053L) and 3) NEB Quick Ligation Module (catalog number E6056L). Each of the modules was diluted 1:2 using the appropriate buffers. Each library was amplified for 12 cycles of PCR using Roche Kapa HiFi polymerase (catalog number KK2612). An 8nt unique index sequence was used on each forward and reverse primer. Perkin-Elmer LabChip Caliper GX was used to measure insert sizes. Roche Kapa SYBR FAST Universal kit (catalog number KR0389) was used to quantify libraries which were diluted to 2nM final stocks, pooled in equal molar amounts and sequenced on the Illumina NovaSeq 6000 platform. Samples were sequenced to a depth of 100M paired reads (median: 115,498,752, IQR: 27,788,080) at 125 nucleotide read lengths. Demultiplexing was based on the unique i5 and i7 indexes to individual sample FASTQ files.

### Genome and Transcriptome Alignment and Quantification

STAR v2.6.1d^6^ was used to align FASTQs to the human genome (B38). Gene counts were generated with featureCounts v1.6.2 using the GENCODE29 annotation^7^. Transcripts were estimated with salmon v0.11.3 from fastq files using GENCODE29^8^. Table 2 describes the number of genes and transcripts detected across the cohort.

**Table 2.** Gene and transcript diversity. A whole transcriptome, strand-specific library prep (as opposed to a polyA library prep) was performed; therefore, we detected a number of lncRNA and other non-coding RNA genes and transcripts in addition to protein-coding genes and transcripts. The mean number of genes detected at one count or more from featureCounts and transcripts detected at a TPM greater than zero are reported, with one standard deviation.

| Biotype | PPMI Genes | PPMI Transcripts |
| --- | --- | --- |
| Protein coding | 17,279 ± 1,312 | 58,473 ± 3,561 |
| lncRNA | 9,123 ± 2,674 | 54,866 ± 5,541 |
| ncRNA | 2,617 ± 931 | 2,789 ± 672 |
| Pseudogene | 6,832 ± 3,009 | 5,711 ± 2,985 |
| Other | 1,017 ± 183 | 1,209 ± 121 |
| Total | 36,868 | 123,048 |

### Cell Count Data

Blood cell populations contribute significantly to the genes detected in whole blood samples. Complete blood count with differential (CBC with diff) was collected for many of the samples in the PPMI study. 1999 of the samples in our study had a corresponding CBC and this can be accessed through LONI. Samples taken at baseline do not have cell count data available.

### Data Records

Raw sequencing data (fastqs), alignment files (BAMs), TPMs and counts for each sample are available through the LONI Image Data Archive (IDA). (FAIRsharing.org: IDA; LONI Image Data Archive; DOI: https://doi.org/10.25504/FAIRsharing.r4ph5f). Additional data, including but not limited to study arm, motor assessments, DaTSCAN and MRI imaging, genetic testing results, whole exome and genome sequencing data, patient history, and standardized techniques and protocols for data collection are also available through the IDA. To access the complete data, researchers need to fill out a Data Use Agreement.

### Technical Validation

#### Assessment of Inter-plate Variability

In order to reduce variability, RNA from whole blood samples was sent to HAIB for library preparation using automated protocols and similar lot numbers throughout sequencing. Libraries were generated across fifty-three 96-well plates for sequencing. Insert sizes can influence the number of genes detected in a sample, and are displayed in Figure 2B for each plate. Insert size by plate accounts for some of the greatest technical variability in this study. There were 53 total plates that were sequenced in two phases of the project, plates 001-028 being the first phase and plates 201-225 being the second phase. As an example of the variance, inserts run on plates 1 and 220 contain larger insert sizes where plate 1 is the largest outlier. Plate 3 and plate 15 have smaller inserts, coinciding with lower numbers of genes detected for samples on those plates (Figure 2b). By design, the baseline, 1-, 2- and 3-year samples for each individual were kept on the same plate if possible to reduce intra-subject variability.

#### Sample Library Quality Control (QC)

RNA sequencing samples were analysed by a number of QC metrics. Samples with fewer than 50 million paired-end reads were removed. Additionally, outliers were identified with: 1) sex incompatibility checks, check between gene expression determined sex and clinically reported sex, using gene expression data of six sex chromosome genes: XIST, RPS4Y1, RPS4Y2, KDM5D, DDX3Y, and USP97^10^. From this set of genes, samples were clustered using t-Distributed Stochastic Neighbour Embedding (t-SNE) (Figure 3A); 2) a Rosner outlier test, to assess outliers, a principal component analysis (PCA) was performed on the top 20,000 genes based on variance stabilized and transformed count data (DESeq2)^9^, alpha = 0.05/number of samples was performed on PC1 (Figure 3B). Additionally, samples showing discordance between SNP calls from the clinical genotypes and RNA-were excluded. Briefly, a set of 1000 SNPs with expression greater than 20x in 99% of RNA-seq samples, within HWE (<0.01) were compared between pileup calls made with samtools BCFtools and the WGS VCF calls^11^. For each sample, the PI_HAT was used to estimate concordance, where samples less than 0.8 were flagged^12^. A small number of samples did not have WGS, and for these, clinical genotypes for GBA, LRRK2, and SNCA were calculated using PLINK^12^. After the removal of samples failing QC, the final dataset comprised of 4,756 samples from 1,570 subjects (Table 1).

#### Reference Pools

In addition to patient samples, each 96-well plate included the same two pools in order to assess technical variability across plates. The pools reflect technical variability across the project; PD Pool and HC Pool on Plate 1, with the largest insert size, can be seen as outliers (Figure 3C). Gene expression across the pools is highly correlated (Figure 3C; HC pool Spearman’s Rank = 0.991, PD pool Spearman’s Rank = 0.990). The most variable genes, defined as those contributing to more than a sum of 0.1% of variation in the first 10 principle components of each respective PCA, were filtered out for downstream analysis, amounting to the removal of 2,667 genes. The correlation of both HC and PD pools across plates was higher when the variable genes were filtered out (HC pool Spearman’s Rank = 0.996, PD pool Spearman’s Rank = 0.995).

#### Gene and Transcript Diversity

With the goal of creating a highly diverse transcriptome that includes both protein coding RNA and long non-coding RNA (lncRNA), total RNA was sequenced in a strand specific manner to a depth of 100 million read pairs per sample. Due to the increased depth of sequencing in the PPMI cohort, a similar number of protein coding genes and transcripts were detected in the whole blood transcriptome as in the poly-A enriched GTEx whole blood transcriptome (a useful, comparable whole blood dataset)^13^. In addition, the whole transcriptome sequencing approach for PPMI provided a large number of lncRNA genes and lncRNA transcripts (Table 2).

#### Variance Partition Analysis

Variance partition analysis uses a mixed linear model at the gene level to quantify variation for traits of interest ^14^. Here, we examine both technical variables (useable bases, site, plate, and plate well position) and biological variables (individual, age, sex, disease group, ethnicity, lymphocyte percentage, and neutrophil percentage). Variance partition analysis revealed that the top sources of gene-level variation in the dataset are individual subjects, usable bases, site, lymphocytes, plate, and neutrophils (Figure 3D). It should be noted that site can contribute both to technical and biological variation, as some sites consisted of subjects primarily enrolled in the genetic registry and genetic cohort (Figure 2A).

#### Gene-Browser

As part of this study, we have also built a web-based browser (https://ppmi-info.org) allowing users the ability to explore quality metrics across the study or examine gene-level expression. The portal requires signing of a DUA to access, provides high level detail, granularity in exploration of clinical parameters, and exploring data as CSV files or tables. TPM are used to display expression levels. For gene queries, the primary display provides a violin/boxplot of “study cohort”, faceted by “visit”, and the user can change these values across a series of pre-joined clinical variables. The browser is built upon a Vega.js JavaScript graphing framework accessing a JSON API, served by MongoDB database. In addition, other data sources are integrated including public data from Uniprot and previously published GWAS expression data^15^.

#### Usage Notes

While the sequencing data is of overall high quality, there are some sources of technical variability of which to be aware while designing downstream analyses, highlighted here. These sources of variability include the overall plate design, differences in insert size across plates, and rRNA and globin depletion. We recommend using the reference pools, which are for assessing quality metrics and removing highest variance genes from analyses.

#### Disease and Genetic Group Assignments

As many of the subjects in PPMI had a recent diagnosis at baseline, it is possible that the PD cohort contains a small number of subjects with other DAT-deficit parkinsonian syndromes such as progressive supranuclear palsy (PSP), multiple system atrophy (MSA) and cortical basal syndrome (CBS). Therefore, investigators reassessed the subject’s primary diagnosis at each visit. Additionally, subjects may have enrolled in the de novo PD or healthy control study arms and as a result of genetic testing during the study, were shown to be positive for *LRRK2, GBA*, or *SNCA* mutations. Conversely subjects could be enrolled in the Genetic Registry or Genetic Cohort without a positive mutation identified for *LRRK2, GBA*, or *SNCA*. Therefore, we created a protocol for reassigning genetic and disease groups for downstream analysis, based on the study arm at enrollment with updates based on genetic testing records and the primary diagnosis records from each visit [DOI: dx.doi.org/10.17504/protocols.io.bb9nir5e]. Genetic mutation status was determined by genetic testing results available from the LONI Image Data Archive (IDA), using WGS variant data where genetic testing results were unavailable. For those subjects who are prodromal at baseline that develop a PD diagnosis, the primary diagnosis remains prodromal with a notation for the visit on which diagnosis changes to PD. There are also some subjects that entered as unaffected in the genetic cohort or registry that have a PD primary diagnosis at a later visit. These group assignments and how they relate to study enrollment are listed in Table 3.

**Table 3.**
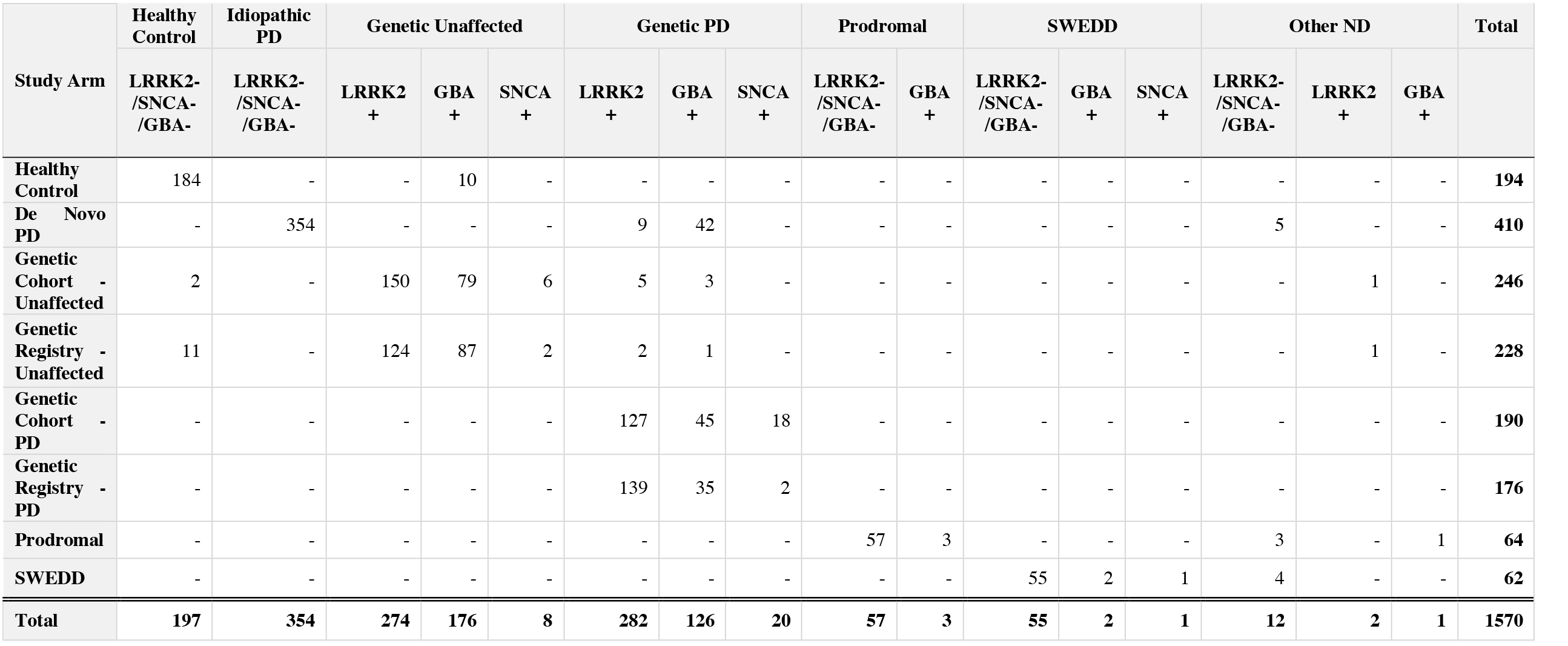
Genetic and Disease Group Assignments. LRRK2+: LRRK2_R1441G OR LRRK2_R1441C OR LRRK2_R1628P OR LRRK2_Y1699C OR LRRK2_G2019S OR LRRK2_G2385R GBA+: GBA_N370S OR GBA_T408M OR GBA_E365K OR GBA_IVS2 OR GBA_84GG OR GBA_L444P SNCA+: SNCA_A53T OR SNCA_E46K OR SNCA_A30P

## Data Availability

Data are available in a public (institutional, general or subject specific) repository that does not issue datasets with DOIs (non-mandated deposition). The sequencing data that support the findings of this study are available from Accelerating Medical Partnerships: Parkinsons Disease (AMP PD; https://amp-pd.org/). These are the requirements for downloading from AMP PD: 1. Personal and institutional/company details; 2. Description of intended data use e.g. proposed analyses; 3. Institutional signature on the AMP PD Data Use Agreement (for researchers requesting access to individual-level, omics data).
All code used for data analysis is available at github: https://github.com/tgen/ppmi-qc-wt-paper

https://www.ppmi-info.org

https://doi.org/10.25504/FAIRsharing.r4ph5f

https://amp-pd.org

## Code Availability

Code used to generate the figures in this paper can be accessed at github (https://github.com/tgen/ppmi-qc-wt-paper).

## Acknowledgements

We thank the Michael J Fox Foundation for funding and providing resources. We thank all of the participants and their families: without their contributions, there would be no study. The samples are collected across many sites, and we thank the many clinical coordinators and administrators that keep these data organized and available to researchers. We thank the groups at Indiana University and HudsonAlpha Institute for Biotechnology for helping to isolate RNA and prepare samples for sequencing, as well as the sequencing. This research was supported and funded by the Michael J Fox Foundation for Parkinson’s Research (MJFF). Many MJFF staff assisted in getting data harmonized and transferred. This research was supported in part by the Intramural Research Program of the National Institute of Health (NIH), National Institute on Aging (NIA). Data used in the preparation of this article were obtained from the PPMI database (www.ppmi-info.org/data). For up-to-date information on the study, visit www.ppmi-info.org. We would also like to acknowledge the industry partners of the PPMI: AbbVie, Allergan, Amathus Therapeutics, Avid Radiopharmaceuticals, Biogen, BioLegend, Bristol Myers Squibb, Celgene, Denali, GE Healthcare, Genentech, GlaxoSmithKline (GSK), Golub Capital, Handl Therapeutics, insitro, Janssen Neuroscience, Lilly, Lundbeck, Merck, Meso Scale Discovery, Neurocrine Biosciences, Pfizer, Piramal, Prevail Therapeutics, Roche, Sanofi Genzyme, Servier, Takeda, Teva, UCB, Verily and Voyager Therapeutics. This research was supported in part by the Intramural Research Program of the National Institute of Health, National Institute on Aging.

## Author contributions

Elizabeth Hutchins; data analyst, study design, writing

David Craig, data analyst, study design, writing

Ivo Violich, data analyst, study design

Eric Alsop, data analyst, study design

Bradford Casey, study design

Samantha Hutten, study design

Alyssa Reimer, study design

Timothy G. Whitsett, study design, writing

Karen L. Crawford, data organization and storage

Artur W, Toga, data organization and storage

Shawn Levy, sequencing

Madison Robinson, sequencing

Nripesh Prasad, sequencing

J. Raphael Gibbs, data analyst, study design

Mark R. Cookson, study design

Kendall Van Keuren-Jensen, data analyst, study design, writing

## Competing interests

BC, SM, AR are employees of the MJFF. EH, DC, IV, ER, AT, KVKJ are funded by MJFF. The authors declare no competing interests.

## Notes

### Competing Interest Statement

BC, SM, AR are employees of the Michael J. Fox Foundation (MJFF). EH, DC, IV, ER, AT, KVKJ are funded by MJFF. The authors declare that the employment or funding from MJFF in no way affected the design, results or interpretations presented.

### Clinical Protocols

http://www.ppmi-info.org

### Funding Statement

This research was supported and funded by the Michael J Fox Foundation for Parkinsons Research (MJFF). Many MJFF staff assisted in getting data harmonized and transferred. This research was supported in part by the Intramural Research Program of the National Institute of Health (NIH), National Institute on Aging (NIA).
BC, SM, AR are employees of the Michael J. Fox Foundation (MJFF). EH, DC, IV, ER, AT, KVKJ are funded by MJFF. The authors declare that the employment or funding from MJFF in no way affected the design, results or interpretations presented.

### Author Declarations

Participants were enrolled at 33 clinical sites across the US (21), Europe (10), Israel (1), and Australia (1). This study was conducted in accordance with the Declaration of Helsinki and the Good Clinical Practice (GCP) guidelines after approval of the local ethics committees of the participating sites. The study was approved by the Research Subjects Review Board at the University of Rochester, and by the institutional review board at each site. Full inclusion and exclusion criteria are listed at clinicaltrials.gov (Identifier: NCT01141023) and at https://ppmi-info.org.

